# Sleep apnoea is a risk factor for severe COVID-19

**DOI:** 10.1101/2020.09.26.20202051

**Authors:** Satu Strausz, Tuomo Kiiskinen, Martin Broberg, Sanni E. Ruotsalainen, Jukka Koskela, Adel Bachour, Finn Gen, Aarno Palotie, Tuula Palotie, Samuli Ripatti, Hanna M. Ollila

## Abstract

**Objective:** To investigate if obstructive sleep apnoea (OSA) is an independent risk factor for severe COVID-19. To examine whether the risk for contracting COVID-19 is elevated among OSA patients.

**Design and setting:** Registry based retrospective case-control study using Finnish nationwide health registries and the FinnGen Study cohort.

**Participants:** Information regarding OSA diagnosis and COVID-19 infection was extracted from the FinnGen study (N=260,405) with a total of 305 patients who had a recorded PCR-validated COVID-19 infection including 26 (8.5%) individuals who were also OSA patients. Severe COVID-19 (N=83; 27.2%) was defined as an infection requiring hospitalization. Among the hospitalized individuals there were 16 (19.3%) with OSA diagnosis. In addition, we also included in our analysis previously reported risk factors for both severe COVID-19 or risk factors and comorbidities for OSA from FinnGen.

**Main outcome measures:** OSA diagnosis, information concerning COVID-19 infection such as hospitalization, were derived from Finnish National Hospital Discharge Registry, Causes of Death Registry and the National Infectious Diseases Registry.

**Results:** We show that OSA is a risk factor for COVID-19 hospitalization independent from age, sex, body mass index (BMI), hypertension, diabetes, coronary heart disease (CHD), asthma and chronic obstructive pulmonary disease (COPD), (p-unadjusted=1.04×10^−4^, OR- adjusted=5.14 [95%CI 1.31 to 22.91], p-adjusted=0.023). OSA was not associated with the risk of contracting COVID-19 (p=0.49).

**Conclusion:** While an OSA patient’s risk of contracting COVID-19 is the same as non-OSA individuals, the OSA patients have a five-fold risk to be hospitalized when affected by COVID-19 than non-OSA individuals. Our findings suggest OSA as one of the risk factors for severe COVID-19. While these associations are statistically significant, they would benefit from replication in an independent cohort.

## Introduction

COVID-19 is a severe respiratory disease caused by SARS-COVID-2 virus infection. A subset of patients face hospitalization, respiratory failure or even death. The severity of COVID-19 is highly age dependent but also evidenced by the number of individuals that receive hospital and intensive care treatment. This was approximately 1-6% of those tested positive for the virus in Finland^1^ and similar to those reported by Centers for Disease Control and Prevention (CDC) ranging from 0.1-5% with current estimates^2, 3^. Severe COVID-19 outcome is mediated primarily through respiratory distress^4, 5^. Risk factors for severe COVID-19 have been identified as older age, male sex, obesity, diabetes, cardiovascular disease and poor lung function^6^. In addition, other respiratory diseases have been listed as potential contributors for COVID-19 severity. Indeed, three small studies have suggested that obstructive sleep apnoea (OSA) may be a risk factor for severe COVID-19^7–9^. Such risk would have substantial effect as OSA is a common disease affecting at least 8% of the population with higher prevalence in older age groups reaching to over 20% in individuals over 60 years of age^10^. The disease etiology of OSA is characterized by repetitive apnoea-hypopnea cycles during sleep causing shortness of breath which is associated with sometime severe oxygen desaturation, sleep disruption and increase in systolic and diastolic blood pressure^11^. The known risk factors for OSA include obesity, high age, male sex and craniofacial and upper-airway structure variations and anomalies^12^. Similarly, OSA is associated with increased risk for cardiovascular mortality, especially if not treated. Finally, it is essential to note that treatment exists for majority of OSA patients so that night-time breathing can be supported by continuous positive airway pressure (CPAP) or mandibular advancement device (MAD). This treatment substantially decreases the risk for cardiovascular events and death^13, 14^.

We set to test the association between earlier OSA diagnosis and transmission risk and severity of COVID-19. We tested the association in the FinnGen study (N=260,405). FinnGen combines major health event data from a) the Finnish National Hospital Discharge Registry, b) Causes of Death Registry and c) Medication Reimbursement Registry and d) the National Infectious Diseases Registry, which provides information about incidence and severity of COVID-19 infection. In addition, demographic and anthropometric data of body mass index (BMI) and smoking status are included.

We specifically aimed at evaluating if OSA associates with the risk for severe COVID-19 infection independently of other potential risk factors including age, sex, BMI, hypertension, diabetes (including type 1 and type 2 diabetes), coronary heart disease (CHD), asthma and chronic obstructive pulmonary disease (COPD), and also whether the risk for contracting COVID-19 is elevated among OSA patients.

## Methods

### Study sample set

FinnGen (https://www.finngen.fi/en) is a large biobank study, aiming to genotype 500,000 Finns, which includes prospective epidemiological and disease-based cohorts, and hospital biobank samples (**Supplementary Table 1**). The data were linked by the unique national personal identification numbers to National Hospital Discharge, Death, and Medication Reimbursement Registries. The Ethics Review Board of the Hospital District of Helsinki and Uusimaa approved the FinnGen study protocol (HUS/990/2017).

The data included of 260,405 Finnish individuals from FinnGen Preparatory Phase Data Freeze 6 with 305 COVID-19 patients. Of them, 26 (8.5%) had OSA. Severe COVID-19 cases (N=83) included 16 (19.3%) OSA patients. Severe COVID-19 was defined as an infection requiring hospitalization. The information of COVID-19 positive individuals was collected from the National Infectious Diseases Registry and the infection was verified by laboratory test using PCR-testing. Data for risk factors, comorbidities and OSA diagnosis was obtained from the Finnish National Hospital Discharge Registry, Causes of Death Registry and Medication Reimbursement Registry, where the clinical endpoints had been generated using International Classification of Diseases (ICD)-codes for OSA, hypertension, diabetes (including type 1 and type 2 diabetes), CHD, asthma and COPD, (**Supplementary Table 2**).

The diagnosis in this special health care derived registry data for OSA is typically based on the following criteria: subjective symptoms, clinical examination and sleep registration applying apnoea-hypopnea-index (AHI) 5 ≥ hour for polysomnography or respiratory event index (REI) 5 ≥ hour for out-of-center sleep study (OCST).

### Patient and public involvement

Patients and public were not involved in the designing process of this study. The patients will not be informed individually of the study results otherwise than through possible media coverage.

### Statistical methods

Differences in baseline demographics and clinical characteristics were tested using on X^2^ tests. Fisher’s exact test was used if the expected cell size was ≤ 5. For continuous variables Student’s t-test was used. We considered p < 0.05 as statistically significant, and all tests were two sided (**Table 1, Table 2**). Logistic regression was used to calculate odds between hospitalized and non-hospitalized groups and similarly between COVID-19-positive and non-COVID-19 groups. The model was adjusted for age, sex, OSA, BMI, hypertension, diabetes (including type 1 and type 2 diabetes), CHD, asthma and COPD, (Table 1). The R statistical package (V.4.0.2) was used for all analyses (www.r-project.org).

**Table 1:**
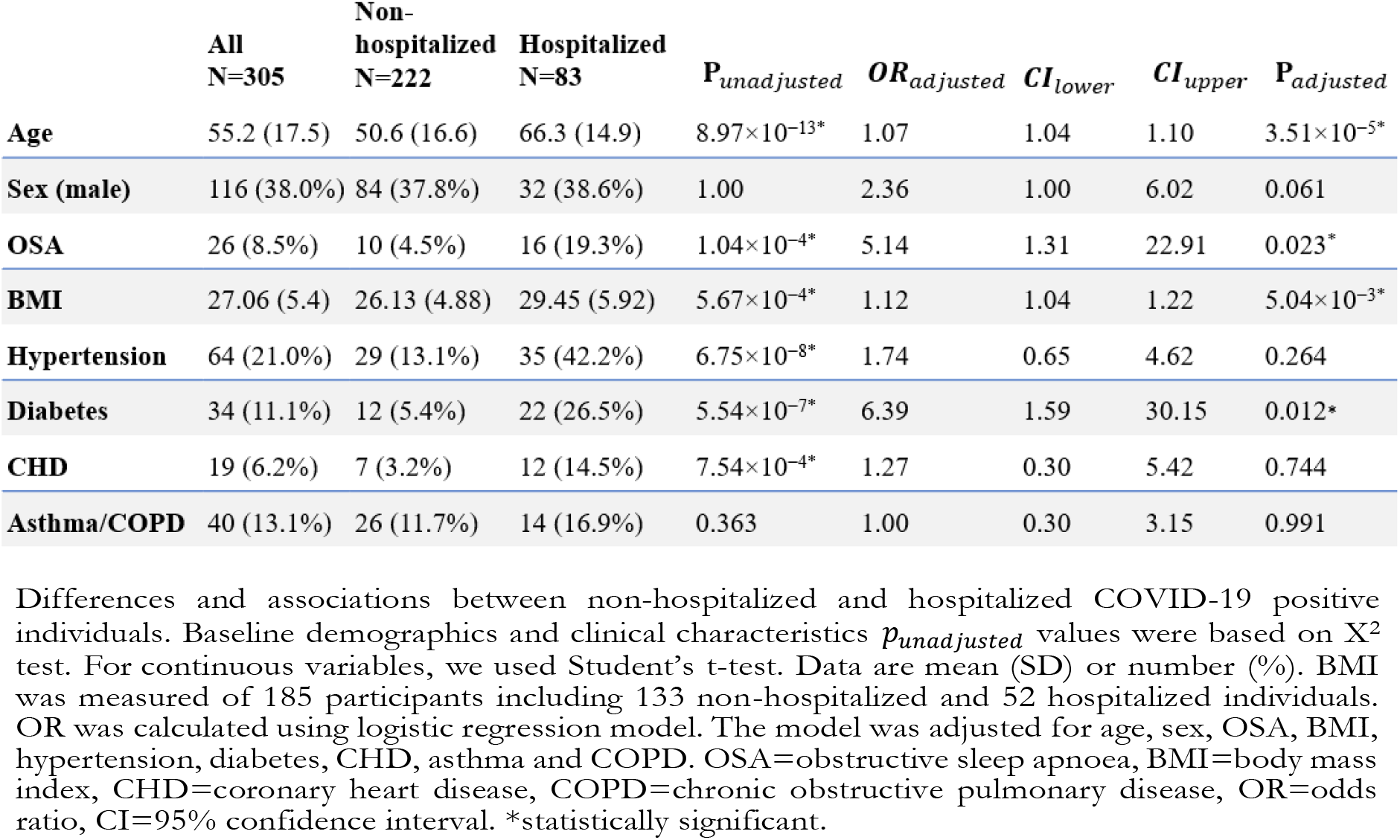
Comparison of the baseline characteristics among COVID-19 positive individuals

**Table 2:**
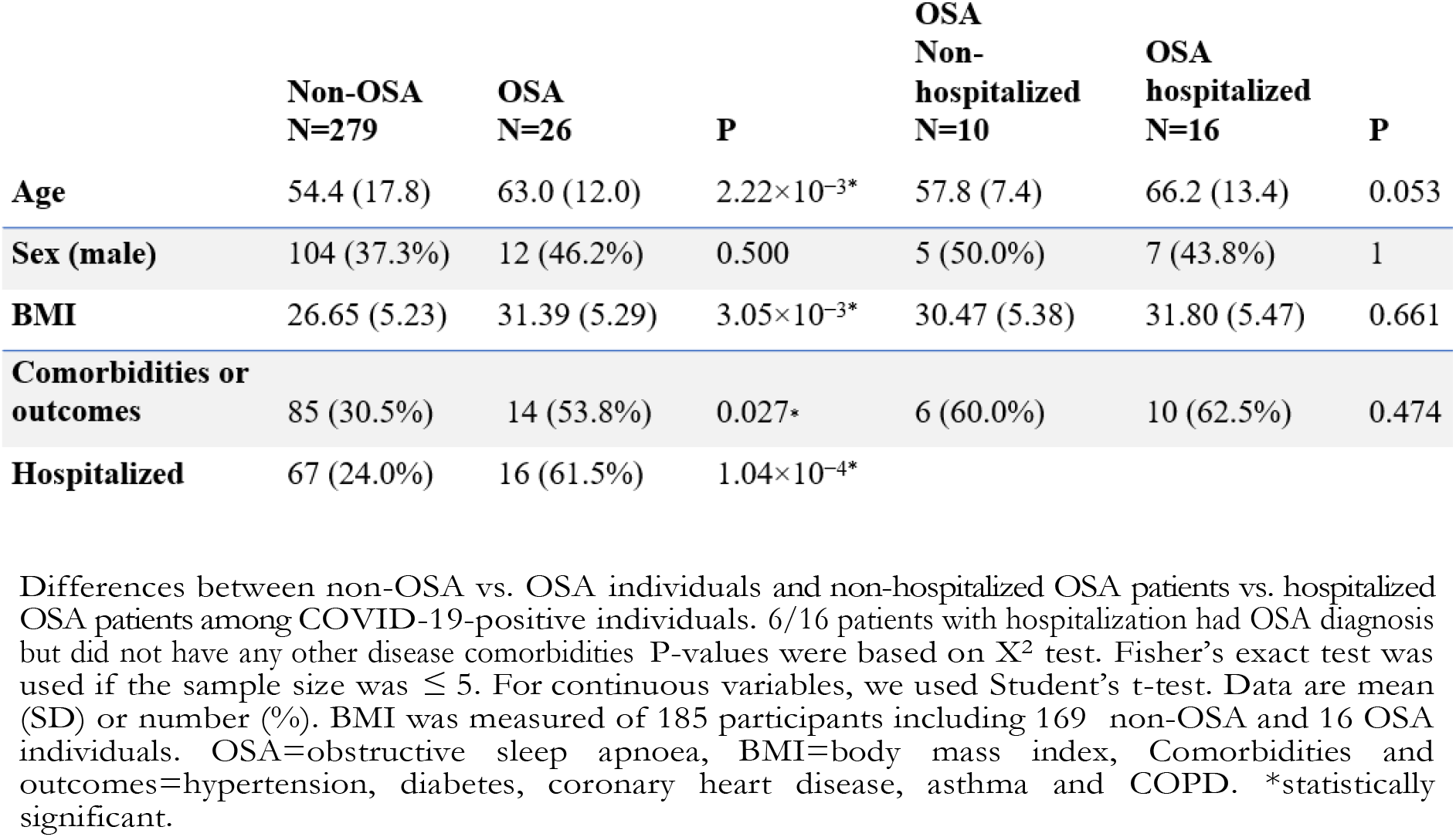
Description of COVID-19 patients with or without OSA and a comparison between non-hospitalized and hospitalized OSA patients

## Results

Of all patients with COVID-19 diagnosis (N=305, 38% male, mean age 55.2 years) 26 patients also had OSA diagnosis (8.5%, 46.2% male, mean age 63.0), (Table 1, Table 2). This reflects a similar prevalence of OSA diagnoses in COVID-19 infected as in the normal population in FinnGen, where prevalence is 8%^15^. 83 patients required hospitalization (38.6% male, mean age 66.3 years) including 16 OSA patients.

Prevalence of OSA (p=1.04×10^−4^), hypertension (p=6.75×10^−8^), diabetes (p=5.54×10^−7^) and CHD (p=7.54×10^−4^) were higher in the hospitalized group. Similarly, age and BMI were higher among hospitalized individuals (p=8.97×10^−13^, p=5.67×10^−4^, respectively), (Table 1).

To evaluate the performance of the diagnostic events, we compared the main risk factors among COVID-19 patients between OSA individuals (N=26) and those who did not have OSA diagnosis (N=279). OSA patients were older and their BMI was higher (mean age 63.0 years, BMI 31.39, p=2.22×10^−3^, p=3.05×10^−3^, respectively) than non-OSA individuals (mean age 54.4 years, BMI 26.65). Also comorbidities, such as hypertension, diabetes, CHD, asthma and COPD were more prevalent among OSA individuals (p=0.027) and they faced hospitalization more often (p=1.04×10^−4^). We did not observe differences in risk factors when comparing non-hospitalized (N=10, male 50%, mean age 57.8) and hospitalized OSA patients (N=16, male 43.8%, mean age 66.2 years). Furthermore, 6/16 patients with OSA were hospitalized due to COVID-19 did not have any other disease comorbidities (Table 2).

We tested if OSA is associated with the risk of severe COVID-19 infection, determined as being hospitalized (N=83), or COVID-19 infection in general (N=305). While OSA did not affect the risk of contracting COVID-19 (p=0.49), OSA patients had a considerably elevated risk for being hospitalized due to severe COVID-19 (OR=5.14, [95% CI 1.31 to 22.91], p=0.023, adjusted for age, sex, BMI, hypertension, diabetes, CHD, asthma and COPD (Table 1). Smoking did not affect the association between OSA and severe COVID-19 (OSA adjusted for comorbidities and smoking OR = 5.01, [95%CI 1.25 to 22.64], p=0.027). Similarly, adjusting for interaction with sex and age did not notably change the estimate for OSA (OR = 4.23, [95% CI 1.09-18.15], p=0.041).

## Discussion

Here we examined if OSA is a risk factor for severe COVID-19 leading to hospitalization. Our analyses reveal a five times higher risk for COVID-19 hospitalization in OSA patients compared to others infected with COVID-19 and the effect is independent from other known risk factors for OSA, or those for severe COVID-19 suggesting that OSA is an independent risk factor for COVID-19.

Our findings are in line with the previous reports which have hypothesized the connection between OSA and COVID-19 as they share a number of comorbidities and risk factors^7–9^.

Building on these small studies we set to test the role of OSA on COVID-19 hospitalization in a large-scale biobank with key potential confounding factors. Using harmonized registry information of both infections and formal ICD-code based diagnoses in FinnGen we were able to establish OSA as an independent risk factor and show that the effect is not modified by overall COVID-19 infection but directly related to COVID-19 severity instead.

In our study OSA patients had five times higher risk of being hospitalized and the estimate was comparable and independent of the risk in diabetes patients, where elevated risk has been reported earlier for severe COVID-19^16^. Furthermore, as OSA acerbates the effects of many underlying risk factors increasing blood pressure and decreasing oxygen saturation level, our findings together with earlier reports suggest that OSA should to be taken into account when assessing who will develop life threatening complications of COVID-19 infection. Previous studies have reported the harmfulness of COVID-19 especially among men^17, 18^. In our study, COVID-19 infection was more prevalent in women than in men. Similarly, women with a severe infection were older than men (69.5 years, 61.9 years, respectively). These characteristics may explain why the majority of the hospitalized individuals were women.

Our findings should be interpreted in the context that registry-based ascertainment through hospitalization may miss non-hospitalized OSA cases (false negatives) and treatment information such as CPAP or MAD compliance. Also, it would be beneficial to replicate these findings in an independent cohort.

## Conclusions

OSA patients have the same risk of contracting COVID-19 than non-OSA individuals. Meanwhile, in this study, OSA patients had a five times higher risk to be hospitalized when affected by COVID-19 than non-OSA individuals. Our findings may suggest that, in assessment of patients with suspected or confirmed COVID-19 infection, OSA should be recognized as one of the comorbidity risk factors for developing a severe form of the disease.

## Data Availability

The FinnGen data may be accessed through Finnish Biobanks FinnBB portal (www.finbb.fi) and THL Biobank data may be accessed through THL Biobank (https://thl.fi/en/web/thl-biobank).

## Footnotes

### Contributors

TK, HMO, SR and SS conceived the study and designed the study protocol. TK, HMO and SS conducted the literature review, statistical analysis and drafted the manuscript. MB and SER contributed statistical analysis and TK phenotyped study samples. AB, MB, JK, SR, SER, AP, TP and SR reviewed the manuscript for intellectual content, made revisions as needed and approved the final version for publication. HMO, TP and SR supervised the study. The corresponding author attests that all listed authors meet authorship criteria and that no others meeting the criteria have been omitted.

### Transparency declaration

The corresponding author affirms that this manuscript is an honest, accurate, and transparent account of the study being reported; that no important aspects of the study have been omitted; and that any discrepancies from the study as planned have been explained.

### Data availability

The FinnGen data may be accessed through Finnish Biobanks’ FinnBB portal (www.finbb.fi) and THL Biobank data may be accessed through THL Biobank (https://thl.fi/en/web/thl-biobank).

## Acknowledgements

We would like to thank all participants of the FinnGen study for their generous participation. Patients and control subjects in FinnGen provided informed consent for biobank research, based on the Finnish Biobank Act. Alternatively, older research cohorts, collected prior the start of FinnGen (in August 2017), were collected based on study-specific consents and later transferred to the Finnish biobanks after approval by Fimea, the National Supervisory Authority for Welfare and Health. Recruitment protocols followed the biobank protocols approved by Fimea. The Coordinating Ethics Committee of the Hospital District of Helsinki and Uusimaa (HUS) approved the FinnGen study protocol Nr HUS/990/2017.

The FinnGen study is approved by Finnish Institute for Health and Welfare (permit numbers: THL/2031/6.02.00/2017, THL/1101/5.05.00/2017, THL/341/6.02.00/2018, THL/2222/6.02.00/2018, THL/283/6.02.00/2019, THL/1721/5.05.00/2019, THL/1524/5.05.00/2020, and THL/2364/14.02/2020), Digital and population data service agency (permit numbers: VRK43431/2017-3, VRK/6909/2018-3, VRK/4415/2019-3), the Social Insurance Institution (permit numbers: KELA 58/522/2017, KELA 131/522/2018, KELA 70/522/2019, KELA 98/522/2019, KELA 138/522/2019, KELA 2/522/2020, KELA 16/522/2020 and Statistics Finland (permit numbers: TK-53-1041-17 and TK-53-90-20).

The Biobank Access Decisions for FinnGen samples and data utilized in FinnGen Data Freeze 6 include: THL Biobank BB2017_55, BB2017_111, BB2018_19, BB_2018_34, BB_2018_67, BB2018_71, BB2019_7, BB2019_8, BB2019_26, BB2020_1, Finnish Red Cross Blood Service Biobank 7.12.2017, Helsinki Biobank HUS/359/2017, Auria Biobank AB17-5154, Biobank Borealis of Northern Finland_2017_1013, Biobank of Eastern Finland 1186/2018, Finnish Clinical Biobank Tampere MH0004, Central Finland Biobank 1-2017, and Terveystalo Biobank STB 2018001.

