## Supplementary Table 1, Supplementary Table 2 for "Sleep apnoea is a risk factor for severe COVID-19"

### Contributors of FinnGen

#### Steering Committee

| | |
|---|---|
| Aarno Palotie | Institute for Molecular Medicine Finland, HiLIFE, University of Helsinki, Finland |
| Mark Daly | Institute for Molecular Medicine Finland, HiLIFE, University of Helsinki, Finland |

**Pharmaceutical companies**

| | |
|---|---|
| Howard Jacob | Abbvie, Chicago, IL, United States |
| Athena Matakidou | Astra Zeneca, Cambridge, United Kingdom |
| Heiko Runz | Biogen, Cambridge, MA, United States |
| Sally John | Biogen, Cambridge, MA, United States |
| Robert Plenge | Celgene, Summit, NJ, United States |
| Mark McCarthy | Genentech, San Francisco, CA, United States |
| Julie Hunkapiller | Genentech, San Francisco, CA, United States |
| Meg Ehm | GlaxoSmithKline, Brentford, United Kingdom |
| Dawn Waterworth | GlaxoSmithKline, Brentford, United Kingdom |
| Caroline Fox | Merck, Kenilworth, NJ, United States |
| Anders Malarstig | Pfizer, New York, NY, United States |
| Kathy Klinger | Sanofi, Paris, France |
| Kathy Call | Sanofi, Paris, France |

**University of Helsinki & Biobanks**

| | |
|---|---|
| Tomi Mäkelä | HiLIFE, University of Helsinki, Finland, Finland |
| Jaakko Kaprio | Institute for Molecular Medicine Finland, HiLIFE, Helsinki, Finland, Finland |
| Petri Virolainen | Auria Biobank / Univ. of Turku / Hospital District of Southwest Finland, Turku, Finland |
| Kari Pulkki | Auria Biobank / Univ. of Turku / Hospital District of Southwest Finland, Turku, Finland |
| Terhi Kilpi | THL Biobank / Finnish Institute for Health and Welfare Helsinki, Finland |
| Markus Perola | THL Biobank / Finnish Institute for Health and Welfare Helsinki, Finland |
| Jukka Partanen | Finnish Red Cross Blood Service / Finnish Hematology Registry and Clinical Biobank, Helsinki, Finland |
| Anne Pitkäranta | Hospital District of Helsinki and Uusimaa, Helsinki, Finland |
| Riitta Kaarteenaho | Northern Finland Biobank Borealis / University of Oulu / Northern Ostrobothnia Hospital District, Oulu, Finland |
| Seppo Vainio | Northern Finland Biobank Borealis / University of Oulu / Northern Ostrobothnia Hospital District, Oulu, Finland |
| Kimmo Savinainen | Finnish Clinical Biobank Tampere / University of Tampere / Pirkanmaa Hospital District, Tampere, Finland |
| Veli-Matti Kosma | Biobank of Eastern Finland / University of Eastern Finland / Northern Savo Hospital District, Kuopio, Finland |
| Urho Kujala | Central Finland Biobank / University of Jyväskylä / Central Finland Health Care District, Jyväskylä, Finland |

**Other Experts/ Non-Voting Members**

| | |
|---|---|
| Outi Tuovila | Business Finland, Helsinki, Finland |
| Minna Hendolin | Business Finland, Helsinki, Finland |
| Raimo Pakkanen | Business Finland, Helsinki, Finland |

#### Scientific Committee

**Pharmaceutical companies**

| | |
|---|---|
| Jeff Waring | Abbvie, Chicago, IL, United States |
| Bridget Riley-Gillis | Abbvie, Chicago, IL, United States |
| Athena Matakidou | Astra Zeneca, Cambridge, United Kingdom |
| Heiko Runz | Biogen, Cambridge, MA, United States |
| Jimmy Liu | Biogen, Cambridge, MA, United States |
| Shameek Biswas | Celgene, Summit, NJ, United States |
| Julie Hunkapiller | Genentech, San Francisco, CA, United States |
| Dawn Waterworth | GlaxoSmithKline, Brentford, United Kingdom |
| Meg Ehm | GlaxoSmithKline, Brentford, United Kingdom |
| Dorothee Diogo | Merck, Kenilworth, NJ, United States |
| Caroline Fox | Merck, Kenilworth, NJ, United States |
| Anders Malarstig | Pfizer, New York, NY, United States |
| Catherine Marshall | Pfizer, New York, NY, United States |
| Xinli Hu | Pfizer, New York, NY, United States |
| Kathy Call | Sanofi, Paris, France |
| Kathy Klinger | Sanofi, Paris, France |
| Matthias Gossel | Sanofi, Paris, France |

**University of Helsinki & Biobanks**

| | |
|---|---|
| Samuli Ripatti | Institute for Molecular Medicine Finland, HiLIFE, University of Helsinki, Helsinki, Finland |
| Johanna Schleutker | Auria Biobank / Univ. of Turku / Hospital District of Southwest Finland, Turku, Finland |
| Markus Perola | THL Biobank / Finnish Institute for Health and Welfare Helsinki, Finland |
| Mikko Arvas | Finnish Red Cross Blood Service / Finnish Hematology Registry and Clinical Biobank, Helsinki, Finland |
| Olli Carpen | Hospital District of Helsinki and Uusimaa, Helsinki, Finland |
| Reetta Hinttala | Northern Finland Biobank Borealis / University of Oulu / Northern Ostrobothnia Hospital District, Oulu, Finland |
| Johannes Kettunen | Northern Finland Biobank Borealis / University of Oulu / Northern Ostrobothnia Hospital District, Oulu, Finland |
| Reijo Laaksonen | Finnish Clinical Biobank Tampere / University of Tampere / Pirkanmaa Hospital District, Tampere, Finland |
| Arto Mannermaa | Biobank of Eastern Finland / University of Eastern Finland / Northern Savo |

| | |
|---|---|
| | Hospital District, Kuopio, Finland |
| Juha Paloneva | Central Finland Biobank / University of Jyväskylä / Central Finland Health Care District, Jyväskylä, Finland |
| Urho Kujala | Central Finland Biobank / University of Jyväskylä / Central Finland Health Care District, Jyväskylä, Finland |

**Other Experts/ Non-Voting Members**

| | |
|---|---|
| Outi Tuovila | Business Finland, Helsinki, Finland |
| Minna Hendolin | Business Finland, Helsinki, Finland |
| Raimo Pakkanen | Business Finland, Helsinki, Finland |

#### Clinical Groups

**Neurology Group**

| | |
|---|---|
| Hilkka Soininen | Northern Savo Hospital District, Kuopio, Finland |
| Valtteri Julkunen | Northern Savo Hospital District, Kuopio, Finland |
| Anne Remes | Northern Ostrobothnia Hospital District, Oulu, Finland |
| Reetta Kälviäinen | Northern Savo Hospital District, Kuopio, Finland |
| Mikko Hiltunen | Northern Savo Hospital District, Kuopio, Finland |
| Jukka Peltola | Pirkanmaa Hospital District, Tampere, Finland |
| Pentti Tienari | Hospital District of Helsinki and Uusimaa, Helsinki, Finland |
| Juha Rinne | Hospital District of Southwest Finland, Turku, Finland |
| Adam Ziemann | Abbvie, Chicago, IL, United States |
| Jeffrey Waring | Abbvie, Chicago, IL, United States |
| Sahar Esmaeeli | Abbvie, Chicago, IL, United States |
| Nizar Smaoui | Abbvie, Chicago, IL, United States |
| Anne Lehtonen | Abbvie, Chicago, IL, United States |
| Susan Eaton | Biogen, Cambridge, MA, United States |
| Heiko Runz | Biogen, Cambridge, MA, United States |
| Sanni Lahdenperä | Biogen, Cambridge, MA, United States |
| Shameek Biswas | Celgene, Summit, NJ, United States |
| John Michon | Genentech, San Francisco, CA, United States |
| Geoff Kerchner | Genentech, San Francisco, CA, United States |
| Julie Hunkapiller | Genentech, San Francisco, CA, United States |
| Natalie Bowers | Genentech, San Francisco, CA, United States |
| Edmond Teng | Genentech, San Francisco, CA, United States |
| John Eicher | Merck, Kenilworth, NJ, United States |
| Vinay Mehta | Merck, Kenilworth, NJ, United States |
| Padhraig Gormley | Merck, Kenilworth, NJ, United States |
| Kari Linden | Pfizer, New York, NY, United States |
| Christopher Whelan | Pfizer, New York, NY, United States |
| Fanli Xu | GlaxoSmithKline, Brentford, United Kingdom |
| David Pulford | GlaxoSmithKline, Brentford, United Kingdom |

**Gastroenterology Group**

| | |
|---|---|
| Martti Färkkilä | Hospital District of Helsinki and Uusimaa, Helsinki, Finland |
| Sampsa Pikkarainen | Hospital District of Helsinki and Uusimaa, Helsinki, Finland |
| Airi Jussila | Pirkanmaa Hospital District, Tampere, Finland |
| Timo Blomster | Northern Ostrobothnia Hospital District, Oulu, Finland |
| Mikko Kiviniemi | Northern Savo Hospital District, Kuopio, Finland |
| Markku Voutilainen | Hospital District of Southwest Finland, Turku, Finland |
| Bob Georgantas | Abbvie, Chicago, IL, United States |
| Graham Heap | Abbvie, Chicago, IL, United States |
| Jeffrey Waring | Abbvie, Chicago, IL, United States |
| Nizar Smaoui | Abbvie, Chicago, IL, United States |
| Fedik Rahimov | Abbvie, Chicago, IL, United States |
| Anne Lehtonen | Abbvie, Chicago, IL, United States |
| Keith Usiskin | Celgene, Summit, NJ, United States |
| Joseph Maranville | Celgene, Summit, NJ, United States |
| Tim Lu | Genentech, San Francisco, CA, United States |
| Natalie Bowers | Genentech, San Francisco, CA, United States |
| Danny Oh | Genentech, San Francisco, CA, United States |
| John Michon | Genentech, San Francisco, CA, United States |
| Vinay Mehta | Merck, Kenilworth, NJ, United States |
| Kirsi Kalpala | Pfizer, New York, NY, United States |
| Melissa Miller | Pfizer, New York, NY, United States |
| Xinli Hu | Pfizer, New York, NY, United States |
| Linda McCarthy | GlaxoSmithKline, Brentford, United Kingdom |

**Rheumatology Group**

| | |
|---|---|
| Kari Eklund | Hospital District of Helsinki and Uusimaa, Helsinki, Finland |
| Antti Palomäki | Hospital District of Southwest Finland, Turku, Finland |
| Pia Isomäki | Pirkanmaa Hospital District, Tampere, Finland |
| Laura Pirilä | Hospital District of Southwest Finland, Turku, Finland |
| Oili Kaipiainen-Seppänen | Northern Savo Hospital District, Kuopio, Finland |
| Johanna Huhtakangas | Northern Ostrobothnia Hospital District, Oulu, Finland |
| Bob Georgantas | Abbvie, Chicago, IL, United States |
| Jeffrey Waring | Abbvie, Chicago, IL, United States |
| Fedik Rahimov | Abbvie, Chicago, IL, United States |
| Apinya Lertratanakul | Abbvie, Chicago, IL, United States |
| Nizar Smaoui | Abbvie, Chicago, IL, United States |
| Anne Lehtonen | Abbvie, Chicago, IL, United States |
| David Close | Astra Zeneca, Cambridge, United Kingdom |
| Marla Hochfeld | Celgene, Summit, NJ, United States |
| Natalie Bowers | Genentech, San Francisco, CA, United States |

John Michon Genentech, San Francisco, CA, United States
Dorothee Diogo Merck, Kenilworth, NJ, United States
Vinay Mehta Merck, Kenilworth, NJ, United States
Kirsi Kalpala Pfizer, New York, NY, United States
Nan Bing Pfizer, New York, NY, United States
Xinli Hu Pfizer, New York, NY, United States
Jorge Esparza Gordillo GlaxoSmithKline, Brentford, United Kingdom
Nina Mars Institute for Molecular Medicine Finland, HiLIFE, University of Helsinki, Helsinki, Finland

**Pulmonology Group**

Tarja Laitinen Pirkanmaa Hospital District, Tampere, Finland
Margit Pelkonen Northern Savo Hospital District, Kuopio, Finland
Paula Kauppi Hospital District of Helsinki and Uusimaa, Helsinki, Finland
Hannu Kankaanranta Pirkanmaa Hospital District, Tampere, Finland
Terttu Harju Northern Ostrobothnia Hospital District, Oulu, Finland
Nizar Smaoui Abbvie, Chicago, IL, United States
David Close Astra Zeneca, Cambridge, United Kingdom

Steven GreenbergCelgene, Summit, NJ, United States
Hubert Chen Genentech, San Francisco, CA, United States
Natalie Bowers Genentech, San Francisco, CA, United States
John Michon Genentech, San Francisco, CA, United States
Vinay Mehta Merck, Kenilworth, NJ, United States
Jo Betts GlaxoSmithKline, Brentford, United Kingdom
Soumitra Ghosh GlaxoSmithKline, Brentford, United Kingdom

**Cardiometabolic Diseases Group**

Veikko Salomaa Finnish Institute for Health and Welfare Helsinki, Finland
Teemu Niiranen Finnish Institute for Health and Welfare Helsinki, Finland
Markus Juonala Hospital District of Southwest Finland, Turku, Finland
Kaj Metsärinne Hospital District of Southwest Finland, Turku, Finland
Mika Kähönen Pirkanmaa Hospital District, Tampere, Finland
Juhani Junttila Northern Ostrobothnia Hospital District, Oulu, Finland
Markku Laakso Northern Savo Hospital District, Kuopio, Finland
Jussi Pihlajamäki Northern Savo Hospital District, Kuopio, Finland
Juha Sinisalo Hospital District of Helsinki and Uusimaa, Helsinki, Finland
Marja-Riitta Taskinen Hospital District of Helsinki and Uusimaa, Helsinki, Finland
Tiinamaija Tuomi Hospital District of Helsinki and Uusimaa, Helsinki, Finland
Jari Laukkanen Central Finland Health Care District, Jyväskylä, Finland
Ben Challis Astra Zeneca, Cambridge, United Kingdom
Andrew Peterson Genentech, San Francisco, CA, United States
Julie Hunkapiller Genentech, San Francisco, CA, United States

| | |
|---|---|
| Natalie Bowers | Genentech, San Francisco, CA, United States |
| John Michon | Genentech, San Francisco, CA, United States |
| Dorothee Diogo | Merck, Kenilworth, NJ, United States |
| Audrey Chu | Merck, Kenilworth, NJ, United States |
| Vinay Mehta | Merck, Kenilworth, NJ, United States |
| Jaakko Parkkinen | Pfizer, New York, NY, United States |
| Melissa Miller | Pfizer, New York, NY, United States |
| Anthony Muslin | Sanofi, Paris, France |
| Dawn Waterworth | GlaxoSmithKline, Brentford, United Kingdom |

**Oncology Group**

| | |
|---|---|
| Heikki Joensuu | Hospital District of Helsinki and Uusimaa, Helsinki, Finland |
| Tuomo Meretoja | Hospital District of Helsinki and Uusimaa, Helsinki, Finland |
| Olli Carpen | Hospital District of Helsinki and Uusimaa, Helsinki, Finland |
| Lauri Aaltonen | Hospital District of Helsinki and Uusimaa, Helsinki, Finland |
| Annika Auranen | Pirkanmaa Hospital District , Tampere, Finland |
| Peeter Karihtala | Northern Ostrobothnia Hospital District, Oulu, Finland |
| Saila Kauppila | Northern Ostrobothnia Hospital District, Oulu, Finland |
| Päivi Auvinen | Northern Savo Hospital District, Kuopio, Finland |
| Klaus Elenius | Hospital District of Southwest Finland, Turku, Finland |
| Relja Popovic | Abbvie, Chicago, IL, United States |
| Jeffrey Waring | Abbvie, Chicago, IL, United States |
| Bridget Riley-Gillis | Abbvie, Chicago, IL, United States |
| Anne Lehtonen | Abbvie, Chicago, IL, United States |
| Athena Matakidou | Astra Zeneca, Cambridge, United Kingdom |
| Jennifer Schutzman | Genentech, San Francisco, CA, United States |
| Julie Hunkapiller | Genentech, San Francisco, CA, United States |
| Natalie Bowers | Genentech, San Francisco, CA, United States |
| John Michon | Genentech, San Francisco, CA, United States |
| Vinay Mehta | Merck, Kenilworth, NJ, United States |
| Andrey Loboda | Merck, Kenilworth, NJ, United States |
| Aparna Chhibber | Merck, Kenilworth, NJ, United States |
| Heli Lehtonen | Pfizer, New York, NY, United States |
| Stefan McDonough | Pfizer, New York, NY, United States |
| Marika Crohns | Sanofi, Paris, France |
| Diptee Kulkarni | GlaxoSmithKline, Brentford, United Kingdom |

**Opthalmology Group**

| | |
|---|---|
| Kai Kaarniranta | Northern Savo Hospital District, Kuopio, Finland |
| Joni Turunen | Hospital District of Helsinki and Uusimaa, Helsinki, Finland |
| Terhi Ollila | Hospital District of Helsinki and Uusimaa, Helsinki, Finland |
| Sanna Seitsonen | Hospital District of Helsinki and Uusimaa, Helsinki, Finland |
| Hannu Uusitalo | Pirkanmaa Hospital District, Tampere, Finland |

| | |
|---|---|
| Vesa Aaltonen | Hospital District of Southwest Finland, Turku, Finland |
| Hannele Uusitalo-Järvinen | Pirkanmaa Hospital District, Tampere, Finland |
| Marja Luodonpää | Northern Ostrobothnia Hospital District, Oulu, Finland |
| Nina Hautala | Northern Ostrobothnia Hospital District, Oulu, Finland |
| Heiko Runz | Biogen, Cambridge, MA, United States |
| Erich Strauss | Genentech, San Francisco, CA, United States |
| Natalie Bowers | Genentech, San Francisco, CA, United States |
| Hao Chen | Genentech, San Francisco, CA, United States |
| John Michon | Genentech, San Francisco, CA, United States |
| Anna Podgornaia | Merck, Kenilworth, NJ, United States |
| Vinay Mehta | Merck, Kenilworth, NJ, United States |
| Dorothee Diogo | Merck, Kenilworth, NJ, United States |
| Joshua Hoffman | GlaxoSmithKline, Brentford, United Kingdom |

**Dermatology Group**

| | |
|---|---|
| Kaisa Tasanen | Northern Ostrobothnia Hospital District, Oulu, Finland |
| Laura Huilaja | Northern Ostrobothnia Hospital District, Oulu, Finland |
| Katariina Hannula-Jouppi | Hospital District of Helsinki and Uusimaa, Helsinki, Finland |
| Teea Salmi | Pirkanmaa Hospital District, Tampere, Finland |
| Sirkku Peltonen | Hospital District of Southwest Finland, Turku, Finland |
| Leena Koulu | Hospital District of Southwest Finland, Turku, Finland |
| Ilkka Harvima | Northern Savo Hospital District, Kuopio, Finland |
| Kirsi Kalpala | Pfizer, New York, NY, United States |
| Ying Wu | Pfizer, New York, NY, United States |
| David Choy | Genentech, San Francisco, CA, United States |
| John Michon | Genentech, San Francisco, CA, United States |
| Nizar Smaoui | Abbvie, Chicago, IL, United States |
| Fedik Rahimov | Abbvie, Chicago, IL, United States |
| Anne Lehtonen | Abbvie, Chicago, IL, United States |
| Dawn Waterworth | GlaxoSmithKline, Brentford, United Kingdom |

#### FinnGen Teams

**Administration Team**

| | |
|---|---|
| Anu Jalanko | Institute for Molecular Medicine Finland, HiLIFE, University of Helsinki, Finland |
| Risto Kajanne | Institute for Molecular Medicine Finland, HiLIFE, University of Helsinki, Finland |
| Ulrike Lyhs | Institute for Molecular Medicine Finland, HiLIFE, University of Helsinki, Finland |

**Communication**

| | |
|---|---|
| Mari Kaunisto | Institute for Molecular Medicine Finland, HiLIFE, University of Helsinki, Finland |

**Analysis Team**


| | |
|---|---|
| Justin Wade Davis | Abbvie, Chicago, IL, United States |
| Bridget Riley-Gillis | Abbvie, Chicago, IL, United States |
| Danjuma Quarless | Abbvie, Chicago, IL, United States |
| Slavé Petrovski | Astra Zeneca, Cambridge, United Kingdom |
| Jimmy Liu | Biogen, Cambridge, MA, United States |
| Chia-Yen Chen | Biogen, Cambridge, MA, United States |
| Paola Bronson | Biogen, Cambridge, MA, United States |
| Robert Yang | Celgene, Summit, NJ, United States |
| Joseph Maranville | Celgene, Summit, NJ, United States |
| Shameek Biswas | Celgene, Summit, NJ, United States |
| Diana Chang | Genentech, San Francisco, CA, United States |
| Julie Hunkapiller | Genentech, San Francisco, CA, United States |
| Tushar Bhangale | Genentech, San Francisco, CA, United States |
| Natalie Bowers | Genentech, San Francisco, CA, United States |
| Dorothee Diogo | Merck, Kenilworth, NJ, United States |
| Emily Holzinger | Merck, Kenilworth, NJ, United States |
| Padhraig Gormley | Merck, Kenilworth, NJ, United States |
| Xulong Wang | Merck, Kenilworth, NJ, United States |
| Xing Chen | Pfizer, New York, NY, United States |
| Åsa Hedman | Pfizer, New York, NY, United States |
| Kirsi Auro | GlaxoSmithKline, Brentford, United Kingdom |
| Clarence Wang | Sanofi, Paris, France |
| Ethan Xu | Sanofi, Paris, France |
| Franck Auge | Sanofi, Paris, France |
| Clement Chatelain | Sanofi, Paris, France |
| Mitja Kurki | Institute for Molecular Medicine Finland, HiLIFE, University of Helsinki, Finland / Broad Institute, Cambridge, MA, United States |
| Samuli Ripatti | Institute for Molecular Medicine Finland, HiLIFE, University of Helsinki, Finland |
| Mark Daly | Institute for Molecular Medicine Finland, HiLIFE, University of Helsinki, Finland |
| Juha Karjalainen | Institute for Molecular Medicine Finland, HiLIFE, University of Helsinki, Finland / Broad Institute, Cambridge, MA, United States |
| Aki Havulinna | Institute for Molecular Medicine Finland, HiLIFE, University of Helsinki, Finland |
| Anu Jalanko | Institute for Molecular Medicine Finland, HiLIFE, University of Helsinki, Finland |
| Kimmo Palin | University of Helsinki, Helsinki, Finland |
| Priit Palta | Institute for Molecular Medicine Finland, HiLIFE, University of Helsinki, Finland |
| Pietro Della Briotta Parolo | Institute for Molecular Medicine Finland, HiLIFE, University of Helsinki, Finland |
| Wei Zhou | Broad Institute, Cambridge, MA, United States |
| Susanna Lemmelä | Institute for Molecular Medicine Finland, HiLIFE, University of Helsinki, Finland |
| Manuel Rivas | University of Stanford, Stanford, CA, United States |

| | |
|---|---|
| Jarmo Harju | Institute for Molecular Medicine Finland, HiLIFE, University of Helsinki, Finland |
| Aarno Palotie | Institute for Molecular Medicine Finland, HiLIFE, University of Helsinki, Finland |
| Arto Lehisto | Institute for Molecular Medicine Finland, HiLIFE, University of Helsinki, Finland |
| Andrea Ganna | Institute for Molecular Medicine Finland, HiLIFE, University of Helsinki, Finland |
| Vincent Llorens | Institute for Molecular Medicine Finland, HiLIFE, University of Helsinki, Finland |
| Antti Karlsson | Auria Biobank / Univ. of Turku / Hospital District of Southwest Finland, Turku, Finland |
| Kati Kristiansson | THL Biobank / Finnish Institute for Health and Welfare Helsinki, Finland |
| Mikko Arvas | Finnish Red Cross Blood Service / Finnish Hematology Registry and Clinical Biobank, Helsinki, Finland |
| Kati Hyvärinen | Finnish Red Cross Blood Service / Finnish Hematology Registry and Clinical Biobank, Helsinki, Finland |
| Jarmo Ritari | Finnish Red Cross Blood Service / Finnish Hematology Registry and Clinical Biobank, Helsinki, Finland |
| Tiina Wahlfors | Finnish Red Cross Blood Service / Finnish Hematology Registry and Clinical Biobank, Helsinki, Finland |
| Miika Koskinen | Hospital District of Helsinki and Uusimaa, Helsinki, Finland BB/HUS/Univ Hosp Districts |
| Olli Carpen | Hospital District of Helsinki and Uusimaa, Helsinki, Finland BB/HUS/Univ Hosp Districts |
| Johannes Kettunen | Northern Finland Biobank Borealis / University of Oulu / Northern Ostrobothnia Hospital District, Oulu, Finland |
| Katri Pylkäs | Northern Finland Biobank Borealis / University of Oulu / Northern Ostrobothnia Hospital District, Oulu, Finland |
| Marita Kalaoja | Northern Finland Biobank Borealis / University of Oulu / Northern Ostrobothnia Hospital District, Oulu, Finland |
| Minna Karjalainen | Northern Finland Biobank Borealis / University of Oulu / Northern Ostrobothnia Hospital District, Oulu, Finland |
| Tuomo Mantere | Northern Finland Biobank Borealis / University of Oulu / Northern Ostrobothnia Hospital District, Oulu, Finland |
| Eeva Kangasniemi | Finnish Clinical Biobank Tampere / University of Tampere / Pirkanmaa Hospital District, Tampere, Finland |
| Sami Heikkinen | Biobank of Eastern Finland / University of Eastern Finland / Northern Savo Hospital District, Kuopio, Finland |
| Arto Mannermaa | Biobank of Eastern Finland / University of Eastern Finland / Northern Savo Hospital District, Kuopio, Finland |
| Eija Laakkonen | Central Finland Biobank / University of Jyväskylä / Central Finland Health Care District, Jyväskylä, Finland |
| Juha Kononen | Central Finland Biobank / University of Jyväskylä / Central Finland Health Care District, Jyväskylä, Finland |

**Sample Collection Coordination**

| | |
|---|---|
| Anu Loukola | Hospital District of Helsinki and Uusimaa, Helsinki, Finland |

**Sample Logistics**

| | |
|---|---|
| Päivi Laiho | THL Biobank / Finnish Institute for Health and Welfare Helsinki, Finland |
| Tuuli Sistonen | THL Biobank / Finnish Institute for Health and Welfare Helsinki, Finland |
| Essi Kaiharju | THL Biobank / Finnish Institute for Health and Welfare Helsinki, Finland |
| Markku Laukkanen | THL Biobank / Finnish Institute for Health and Welfare Helsinki, Finland |
| Elina Järvensivu | THL Biobank / Finnish Institute for Health and Welfare Helsinki, Finland |
| Sini Lähteenmäki | THL Biobank / Finnish Institute for Health and Welfare Helsinki, Finland |
| Lotta Männikkö | THL Biobank / Finnish Institute for Health and Welfare Helsinki, Finland |
| Regis Wong | THL Biobank / Finnish Institute for Health and Welfare Helsinki, Finland |

**Registry Data Operations**

| | |
|---|---|
| Kati Kristiansson | THL Biobank / Finnish Institute for Health and Welfare Helsinki, Finland |
| Hannele Mattsson | THL Biobank / Finnish Institute for Health and Welfare Helsinki, Finland |
| Susanna Lemmelä | Institute for Molecular Medicine Finland, HiLIFE, University of Helsinki, Finland |
| Tero Hiekkalinna | THL Biobank / Finnish Institute for Health and Welfare Helsinki, Finland |

Manuel González Jiménez. THL Biobank / Finnish Institute for Health and Welfare Helsinki, Finland

**Genotyping**

| | |
|---|---|
| Kati Donner | Institute for Molecular Medicine Finland, HiLIFE, University of Helsinki, Finland |

**Sequencing Informatics**

| | |
|---|---|
| Priit Palta | Institute for Molecular Medicine Finland, HiLIFE, University of Helsinki, Finland |
| Kalle Pärn | Institute for Molecular Medicine Finland, HiLIFE, University of Helsinki, Finland |
| Javier Nunez-Fontarnau | Institute for Molecular Medicine Finland, HiLIFE, University of Helsinki, Finland |

**Data Management and IT Infrastructure**

| | |
|---|---|
| Jarmo Harju | Institute for Molecular Medicine Finland, HiLIFE, University of Helsinki, Finland |
| Elina Kilpeläinen | Institute for Molecular Medicine Finland, HiLIFE, University of Helsinki, Finland |
| Timo P. Sipilä | Institute for Molecular Medicine Finland, HiLIFE, University of Helsinki, Finland |
| Georg Brein | Institute for Molecular Medicine Finland, HiLIFE, University of Helsinki, Finland |
| Alexander Dada | Institute for Molecular Medicine Finland, HiLIFE, University of Helsinki, Finland |
| Ghazal Awaisa | Institute for Molecular Medicine Finland, HiLIFE, University of Helsinki, Finland |
| Anastasia Shcherban | Institute for Molecular Medicine Finland, HiLIFE, University of Helsinki, Finland |
| Tuomas Sipilä | Institute for Molecular Medicine Finland, HiLIFE, University of Helsinki, Finland |

**Clinical Endpoint Development**

| | |
|---|---|
| Hannele Laivuori | Institute for Molecular Medicine Finland, HiLIFE, University of Helsinki, Finland |
| Aki Havulinna | Institute for Molecular Medicine Finland, HiLIFE, University of Helsinki, Finland |
| Susanna Lemmelä | Institute for Molecular Medicine Finland, HiLIFE, University of Helsinki, Finland |
| Tuomo Kiiskinen | Institute for Molecular Medicine Finland, HiLIFE, University of Helsinki, Finland |

**Trajectory Team**

| | |
|---|---|
| Tarja Laitinen | Pirkanmaa Hospital District, Tampere, Finland |
| Harri Siirtola | University of Tampere, Tampere, Finland |
| Javier Gracia Tabuenca | University of Tampere, Tampere, Finland |

**Biobank Directors**

| | |
|---|---|
| Lila Kallio | Auria Biobank, Turku, Finland |
| Sirpa Soini | THL Biobank, Helsinki, Finland |
| Jukka Partanen | Blood Service Biobank, Helsinki, Finland |
| Kimmo Pitkänen | Helsinki Biobank, Helsinki, Finland |
| Seppo Vainio | Northern Finland Biobank Borealis, Oulu, Finland |
| Kimmo Savinainen | Tampere Biobank, Tampere, Finland |
| Veli-Matti Kosma | Biobank of Eastern Finland, Kuopio, Finland |
| Teijo Kuopio | Central Finland Biobank, Jyväskylä, Finland |

Supplementary Table 1. The prospective epidemiological and disease-based cohorts, and hospital biobank samples in FinnGen Data Freeze 6

| **Cohort** | **N** |
|---|---|
| Auria Biobank | 29201 |
| Biobank of Central Finland | 7743 |
| Biobank of Eastern Finland | 7765 |
| Blood Service Biobank | 28901 |
| Borealis Biobank | 7390 |
| Biobank Botnia | 8681 |
| Biobank Corogene | 4689 |
| Biobank FinHealth | 5928 |
| Helsinki Biobank | 58693 |
| Tampere Biobank | 12394 |
| Terveystalo Biobank | 1709 |
| THL Biobank ATBC | 3836 |
| THL Biobank FinIPF | 201 |
| THL Biobank FINRISK 1992 | 4931 |
| THL Biobank FINRISK 1997 | 6997 |
| THL Biobank FINRISK 2002 | 6869 |
| THL Biobank FINRISK 2007 | 5143 |
| THL Biobank FINRISK 2012 | 5233 |
| THL Biobank GENERISK | 6898 |
| THL Biobank Health 2000 | 6529 |
| THL Biobank Health 2011 | 708 |
| THL Biobank HHS | 2113 |
| THL Biobank Kuusamo | 123 |
| THL Biobank Migraine | 7717 |
| THL Biobank SUPER | 8466 |
| THL Biobank Diabetes | 10145 |
| THL Biobank Twins | 11402 |
| **Total:** | 260405 |

Supplementary Table 2. ICD-codes for OSA and comorbidities

| | ICD-10 | ICD-9 | ICD-8 |
|---|---|---|---|
| **OSA** | G47.3 | 3472 | |
| **HYPERTENSION** | I10-I13, I15, I67.4 | 4019X, 4029A, 4029B, 4039A, 4040A, 4059A, 4059B, 4372A, 4059X | 400, 401, 402, 403, 404 |
| **DIABETES** | E10-E14 | 250 | 250 |
| **CHD** | I20.0, I21, I22 | 410, 4110 | 410, 411,0 |
| **ASTHMA** | J45, J46 | 493 | 493 |
| **COPD** | J43, J44 | 492, 4912 | 492, 491,04 |

By combining codes from different registries, we generate phenotype "endpoints". Finnish national version for each ICD-codes were used. These ICD-code criteria are all so-called regular expressions. OSA=obstructive sleep apnoea, CHD=coronary heart disease, COPD= chronic obstructive pulmonary disease.
